# Predicting Unplanned Hospital Readmissions in People with Multiple Long-Term Conditions

**DOI:** 10.64898/2026.08.01.26359453

**Authors:** Rebeen Ali Hamad, Alisha Angdembe, Amaani B. Hussain, John Casement, Wasim A. Iqbal, Christian Atallah, Dexter Canoy, Rafael Henkin, David Taylor, Susan Mountain, Michael R. Barnes, Paolo Missier, Nick J. Reynolds, The AI-MULTIPLY Consortium

## Abstract

The prevalence of multiple long-term conditions (MLTCs) is associated with increased healthcare utilisation and an elevated risk of unplanned 30-day hospital readmission. Existing prediction tools predominantly focus on single-disease cohorts and fail to capture the clinical heterogeneity, polypharmacy, and care complexity characteristic of MLTC populations. Using data from 99,207 UK Biobank (UKBB) participants with MLTCs (***≥***2 long-term conditions), we developed Self-HR, a two-stage self-supervised learning framework that learns transferable patient representations from longitudinal clinical data encompassing hospital admission diagnoses, primary care prescriptions, long-term condition histories, and demographic factors. Self-HR achieved an AUROC of 0.92 and AUPRC of 0.75 in the UKBB discovery cohort, outperforming all supervised baselines — including XGBoost, Random Forest, and fully supervised neural networks — across both overall and minority-class metrics. Performance was sustained upon external validation in 79,224 multimorbid individuals from the Clinical Practice Research Datalink (CPRD; AUROC 0.86, F1 score 0.67 for the readmitted class). Self-HR demonstrated superior robustness to partial outcome labelling and class imbalance, maintaining an F1 score of 0.62 for readmitted patients when trained on only 50% labelled data, compared with 0.28 for the best supervised comparator. Ablation analyses identified incident admission diagnoses as the strongest predictive feature, followed by primary care prescriptions and long-term condition history. Beyond binary classification, Self-HR generalised to regression tasks — predicting incident and emergency admission durations — through fine-tuning alone, achieving the lowest MAE and RMSE across all tasks without repeat pretraining. These findings support Self-HR as a data-efficient and generalisable framework for readmission risk prediction in multimorbid populations, with potential to inform proactive discharge planning and targeted post-discharge care.

## 1 Introduction

Patients with multiple long-term conditions (MLTCs) represent a rapidly expanding and clinically vulnerable population. Rising life expectancy, the growing burden of chronic disease, and lifestyle factors have led to increasing multimorbidity, which substantially complicates inpatient and post-discharge care [1, 2]. Notably, multimorbidity does not occur exclusively in the elderly; more than 50% of multimorbid patients develop two or more long-term conditions (LTCs) before 65 years of age [1]. Among these patients, unplanned hospital readmissions (HRs) are common, costly, and frequently preventable [3]. Readmissions are associated with worse clinical outcomes, increased morbidity, psychological distress, and significant strain on healthcare resources [3]. Despite advances in medical and community care, 30-day readmission rates remain persistently high, reported at approximately 20% in the USA in 2019 and increasing in the UK from 12.5% in 2014 to 15.5% in 2021 [2, 4, 5]. Reducing avoidable HRs therefore remains a major clinical and policy priority.

HR reflects a complex interaction between unresolved illness, multimorbidity, polypharmacy, inadequate discharge planning, and fragmented post-discharge care. Readmission risk is shaped by a constellation of factors including comorbidity burden, medication complexity, socioeconomic deprivation, and ethnicity [6]. The financial impact is substantial: unplanned HRs account for approximately 19% of emergency admission costs in the UK National Health Service (NHS) and an estimated $20 billion USD annually in the United States [7, 8]. Importantly, up to 48% of readmissions are considered potentially preventable, highlighting the urgent need for accurate and clinically actionable risk prediction tools [9].

Early identification of high-risk patients at discharge enables targeted interventions such as medication reconciliation, enhanced follow-up, and community care support, which may reduce readmission risk and improve outcomes. Consequently, a large body of research has applied statistical and machine learning (ML) methods to HR prediction using electronic health record (EHR) data [10–12]. Common predictors include comorbidity indices, length of stay, prior admissions, polypharmacy, specific diagnoses, and laboratory results. However, recent systematic reviews report moderate predictive performance, high risk of bias, and poor generalisability across populations [11]. Most existing models rely on predefined features and fully labelled datasets, and critically, few have been developed or validated specifically for patients with MLTCs. This represents a major gap given the clinical complexity, heterogeneity, and growing prevalence of multimorbidity, particularly in patients with early-onset MLTCs.

In this study, we develop Self-HR, a self-supervised learning (SSL) framework for predicting unplanned HR in patients with MLTCs (defined as *≥*2 LTCs) using EHR data from the UK Biobank (UKBB) and the Clinical Practice Research Datalink (CPRD). We use clinical information available at the index admission — demographics, prescriptions, long-term conditions, and admission diagnoses — to generate a 30-day unplanned readmission risk estimate. Unlike conventional supervised models, SSL leverages large volumes of EHR data without requiring readmission labels during pretraining. The model instead learns latent representations, or embeddings, that capture complex interactions between diagnoses, medications, and longitudinal health trajectories. These embeddings are subsequently fine-tuned using a smaller labelled dataset of readmission outcomes, reducing dependence on exhaustive annotation while improving performance in imbalanced and heterogeneous clinical settings [13].

We evaluate the ability of Self-HR to predict unplanned HR across two large, independent multimorbid cohorts and assess whether learned representations generalise to related downstream clinical tasks without task-specific re-engineering. We benchmark Self-HR against conventional ML approaches, examine its robustness to class imbalance and limited outcome labelling, and perform ablation analyses to support clinical interpretability. By developing and externally validating a framework that achieves strong and transferable performance, this study provides a clinically scalable approach to AI-driven risk stratification in multimorbid populations. Our findings support the translation of SSL-based models into discharge workflows, enabling earlier identification of high-risk patients, more proactive post-discharge care, and reduction of preventable readmissions.

## 2 Materials and Methods

### 2.1 Discovery Dataset: UK Biobank

The discovery cohort comprised multimorbid patients aged 40–69 years at enrolment in the UK Biobank (UKBB) study. Data were extracted in March 2021 (application 69836) in compliance with UKBB general ethical approval and following written informed consent from all participants [14]. Extracted data included demographic information collected at enrolment, linked to primary and secondary care electronic health records (EHRs), including Hospital Episode Statistics (HES) data [15]. Primary care records provided clinical diagnoses and prescriptions from the two years preceding each admission event. Clinical diagnoses were encoded using the Read v2 and Read v3 standard disease taxonomies [16].

Participants were eligible for inclusion (Figure 1) if they met all three of the following criteria: (i) linked primary and secondary care records, including hospital readmission data; (ii) at least one inpatient hospital encounter of one or more days, with over 30 days of follow-up to enable detection of readmission within 30 days [17]; and (iii) a primary care timeline confirming multimorbidity, defined as the presence of at least two LTCs. Participants who withdrew consent after data extraction were excluded from all analyses.

**Fig. 1:**
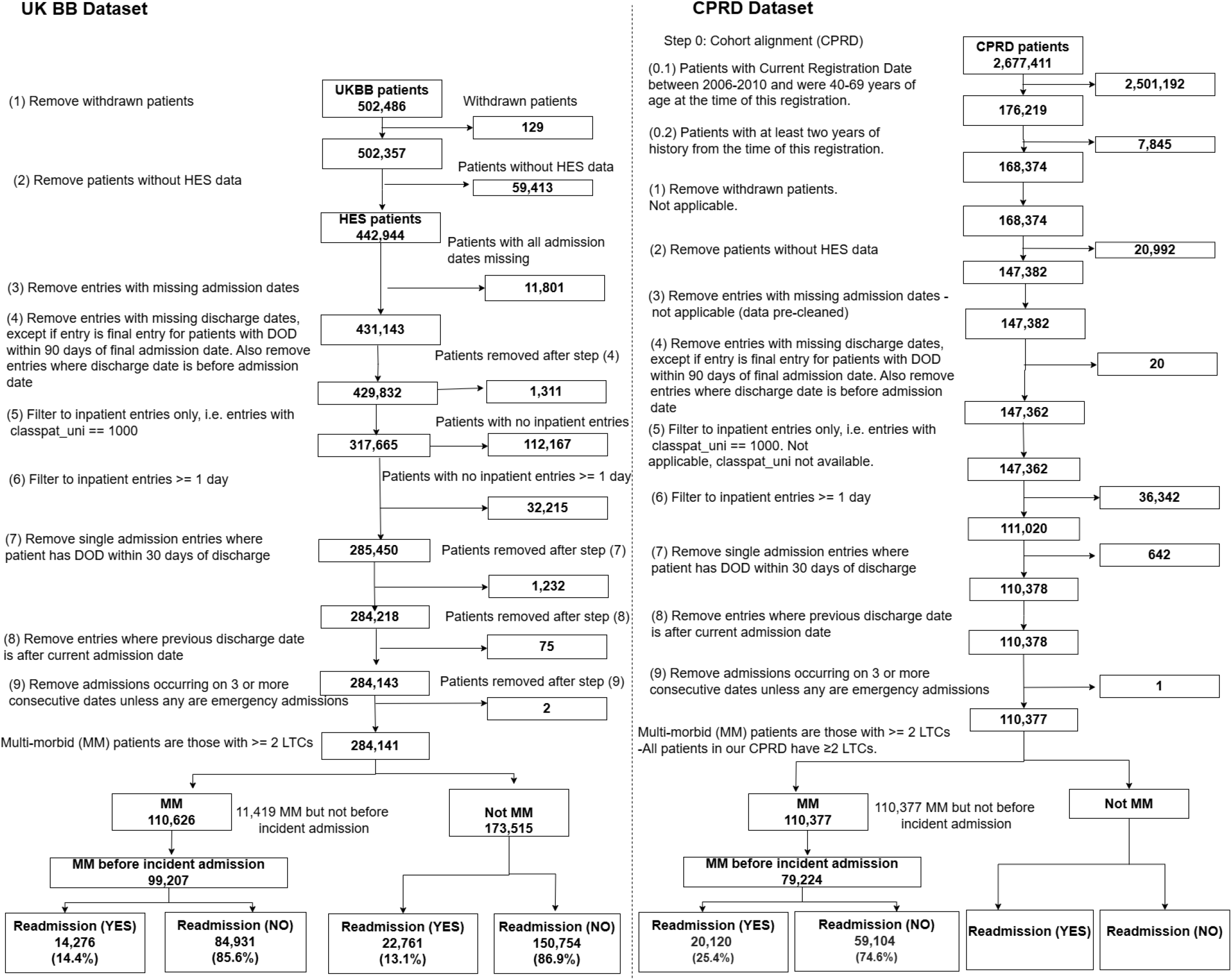
Extracting Readmission datasets from the UKBB and CPRD datasets. CPRD: Clinical Practice Research Datalink, DOD: Date of Discharge, HES: Hospital Episode Statistics, LTC: Long Term Condition, MM: Multimorbid, UKBB: UK Biobank.

Each participant was characterised using four feature groups:

- **Demographic and lifestyle features:** Age, sex, ethnic background, socioeconomic status (Townsend deprivation index), lifestyle factors (alcohol intake frequency, smoking status), and physical health metrics (Body Mass Index (BMI) and body fat percentage).
- **Prescriptions:** Drugs prescribed in the two years prior to the index admission event, classified using the British National Formulary (BNF) at paragraph level [18].
- **Hospital admission diagnoses:** Primary diagnoses recorded at the index hospital admission, classified according to the International Classification of Diseases, 10th revision (ICD-10) [19]. To reduce feature dimensionality and sparsity, ICD-10 codes were ranked by frequency and only those collectively accounting for 90% of all recorded diagnoses across the cohort were retained. Remaining codes were excluded and treated as absent in the one-hot encoded feature space. No hierarchical grouping or aggregation of rare codes was performed.
- **Long-term conditions (LTCs):** A binary indicator for each of the 204 categorised LTCs listed in Supplementary Table 1, identified in GP clinical records using Read v2 codes [16, 20]. Each LTC represents a pre-defined set of clinical terms; for example, the Asbestosis LTC corresponds to the codelist in Supplementary Table 2.

All categorical variables were encoded using one-hot encoding. The LTC feature set was represented as a 204-dimensional binary vector, with a value of 1 indicating a confirmed diagnosis. Following one-hot encoding of all LTCs, prescriptions, and other categorical variables (sex, smoking status, etc.), each participant was represented by a vector of 1,296 features.

### 2.2 Replication Dataset: Clinical Practice Research Datalink (CPRD)

To assess the generalisability of Self-HR, an independent replication cohort was derived from the Clinical Practice Research Datalink (CPRD) GOLD database, which contains anonymised EHRs for individuals registered with general practices in England. Included participants were those aged 40–69 years who registered with a general practice between 2006 and 2010, had two or more LTCs, met CPRD data quality criteria, and had linkage to HES. Additional data were integrated through linkage to the National Death Registry for mortality outcomes and to the Index of Multiple Deprivation (IMD) via the Office for National Statistics (ONS) to capture socioeconomic context. Only patients with at least two years of follow-up data were retained.

Cohort filtering steps were aligned with those applied to the UKBB population (Figure 1), resulting in a final replication cohort of participants with *≥*2 LTCs and at least one hospital admission. To ensure comparability with the UKBB cohort, two variables were excluded from the CPRD analysis: body fat percentage, which was unavailable in CPRD, and alcohol intake frequency, which was excluded due to a high proportion of missing values.

### 2.3 Missing-Data Handling

Missing data were addressed using variable-specific imputation strategies informed by distributional assessments, missingness patterns, and outlier evaluation. Among the ten demographic and lifestyle features, missingness was low overall (all *<*3%). Continuous variables were examined using histograms, skewness metrics, and inter-quartile range (IQR)–based outlier detection. The Townsend deprivation index demonstrated a moderately symmetric distribution with minimal skew and was therefore imputed using the meanBody mass index (BMI) and body fat percentage displayed right-skewed distributions. Outlier detection using IQR-based thresholds identified 2,692 outliers for BMI and 82 for body fat percentage; the substantially higher count for BMI likely reflects its wider population-level variation and greater sensitivity to extreme values at both ends of the distribution. Missing values for both variables were imputed using the median to reduce sensitivity to these extreme observations. Categorical demographic variables—including smoking status, alcohol-drinker status, and ethnic background— were imputed using the mode, preserving the most frequently observed category in each case. Alcohol intake frequency was excluded from the CPRD analysis due to a high proportion of missing values; in the UKBB cohort, where missingness was low, it was retained and imputed using the mode. For prescriptions, diagnoses (ICD-10), and LTCs, missingness reflects the absence of a recorded clinical event rather than incomplete documentation. Consistent with standard practice for EHR–derived binary feature sets, these variables were imputed with zero, indicating that the patient did not receive the corresponding prescription, did not present the specific diagnosis code, or did not have the LTC recorded in primary care data. This combined strategy preserved the structure of the dataset, limited the influence of outliers on imputation, and minimised potential bias associated with low-level missingness.

### 2.4 Outcome definitions

The binary outcome variable, readmission state, defined the negative (not readmitted) and positive (readmitted) prediction classes. Participants were assigned a positive readmission status if they experienced an emergency hospital readmission within 30 days of a previous discharge, identified using admission method codes from the NHS Data Model and Dictionary [21]. These codes correspond to unplanned emergency admissions recorded in Hospital Episode Statistics (HES). Participants without an emergency readmission recorded within this period were assigned a negative readmission status. HES captures all NHS-funded hospital activity in England, including emergency admissions occurring at hospitals different from the site of the original discharge. Therefore, emergency readmissions are expected to be comprehensively captured within the dataset, regardless of geographic location within England. In this study, only emergency admissions, as defined by NHS admission method codes, were considered positive readmissions. Other unplanned but non-emergency admissions (e.g. urgent elective admissions) were not classified as emergency readmissions and were therefore not included in the readmission outcome.

To evaluate the cross-task generalisability of Self-HR, we additionally explored regression-based prediction of admission duration outcomes. Accurate prediction of admission duration is clinically important for bed capacity planning, discharge coordination, and early identification of patients at risk of prolonged stays — particularly in MLTC populations where care complexity frequently drives extended hospitalisations. These tasks were addressed through fine-tuning of the pretrained Self-HR encoder alone, without repeat pretraining, across the following outcomes:

- Emergency readmission: An emergency admission occurring within 30 days of a previous discharge.
- Incident admission: The index hospital admission used as the reference point for readmission assessment. For patients with a subsequent emergency readmission (positive class), this is the preceding discharge from which the 30-day readmission window is measured. For patients without a readmission (negative class), this is their first or only hospital admission after becoming multimorbid.
- First emergency admission: The patient’s first emergency admission after becoming multimorbid.
- Duration: The time difference in days between the admission date and the discharge date.

Figure 2 presents an overview of the proposed framework, where large-scale health records from UK Biobank and CPRD are transformed into multi-modal representations using self-supervised learning to predict unplanned hospital readmission and a set of clinically relevant admission duration outcomes.

**Fig. 2:**
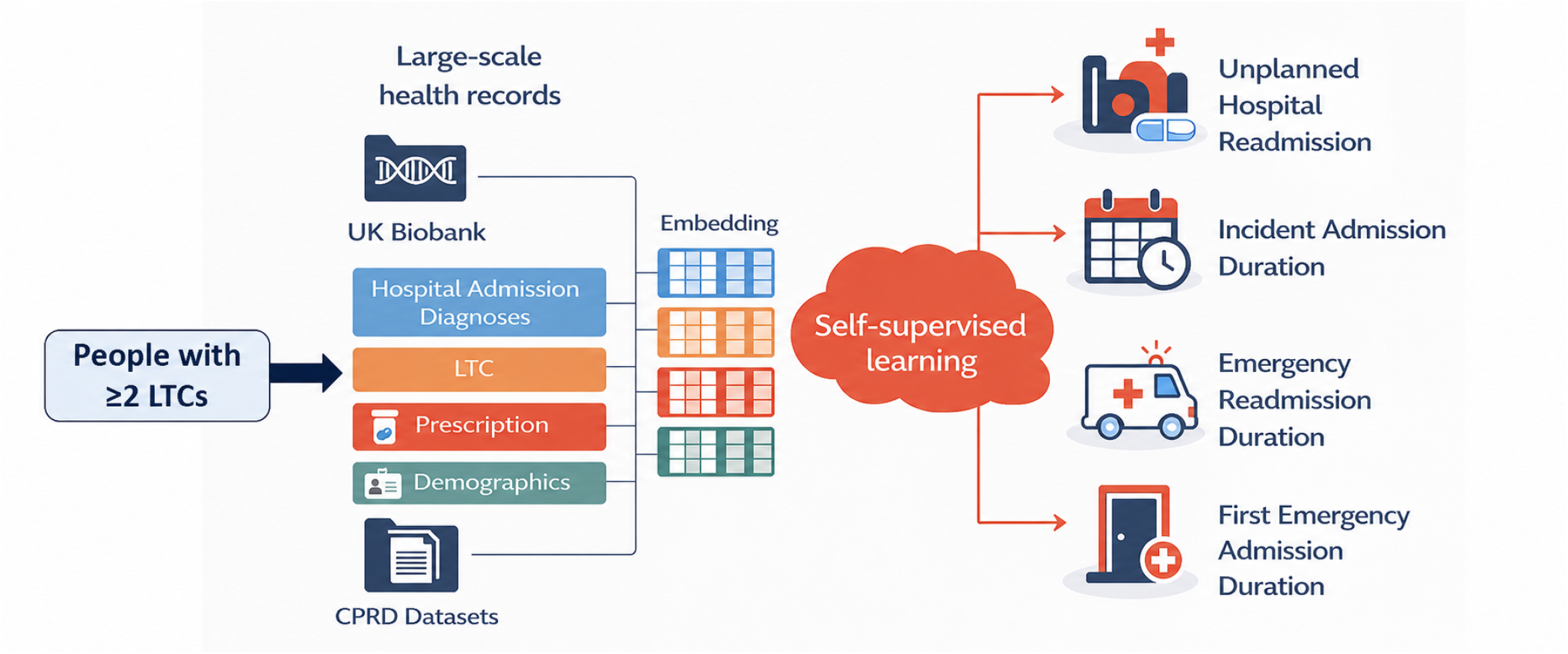
Overview of the proposed self-supervised learning framework leveraging multimodal health records from UK Biobank and CPRD to predict hospital readmission and admission duration outcomes

### 2.5 Addressing class imbalance

The dataset exhibited a pronounced class imbalance, with approximately 5.95 non-readmitted individuals for every one readmitted individual (84,931 vs. 14,276 cases) for UKBB. The corresponding ratio for CPRD was approximately 2.94:1 (59,104 non-readmitted vs. 20,120 readmitted individuals), reflecting a comparatively less severe but still clinically meaningful imbalance.

To address this, we implemented multiple strategies commonly used in ML to mitigate class imbalance, including:

- Random undersampling to match the size of the majority class to the minority class,
- Synthetic Minority Over-sampling Technique (SMOTE) to generate synthetic samples for the minority class,
- Cost-sensitive learning (CSL), where higher penalties are assigned for misclassifying the minority class.

These techniques were used to create various training setups, and the models were evaluated under consistent testing conditions. The two main evaluation strategies were:

1. Baseline evaluation on the original imbalanced dataset using an 80/20 random training/testing split, preserving the natural distribution of readmitted and non-readmitted individuals. This setup served as a baseline to observe how models perform under real-world class distributions.
2. Balanced training using different resampling techniques as above, with stratified testing.

All models trained using these balanced or weighted approaches were evaluated on 20 shuffled stratified test sets that retained the original class distribution. Stratification was performed with respect to the readmission outcome, ensuring that each test set preserved the same proportion of positive and negative cases as the full cohort. For each evaluation run, the data were randomly shuffled prior to stratified splitting to reduce sampling bias. Model performance was then assessed independently on each test set, and we report the mean and standard deviation (SD) of the performance metrics across the 20 test sets to provide a robust estimate under realistic class-imbalance conditions.

### 2.6 Self-supervised Learning (SSL) Model

SSL consists of a *pre-training* phase in which the model uncovers meaningful patterns and representations from the intrinsic structure of the input data, independent of outcome labels (HR status) (Figure 3).

**Fig. 3:**
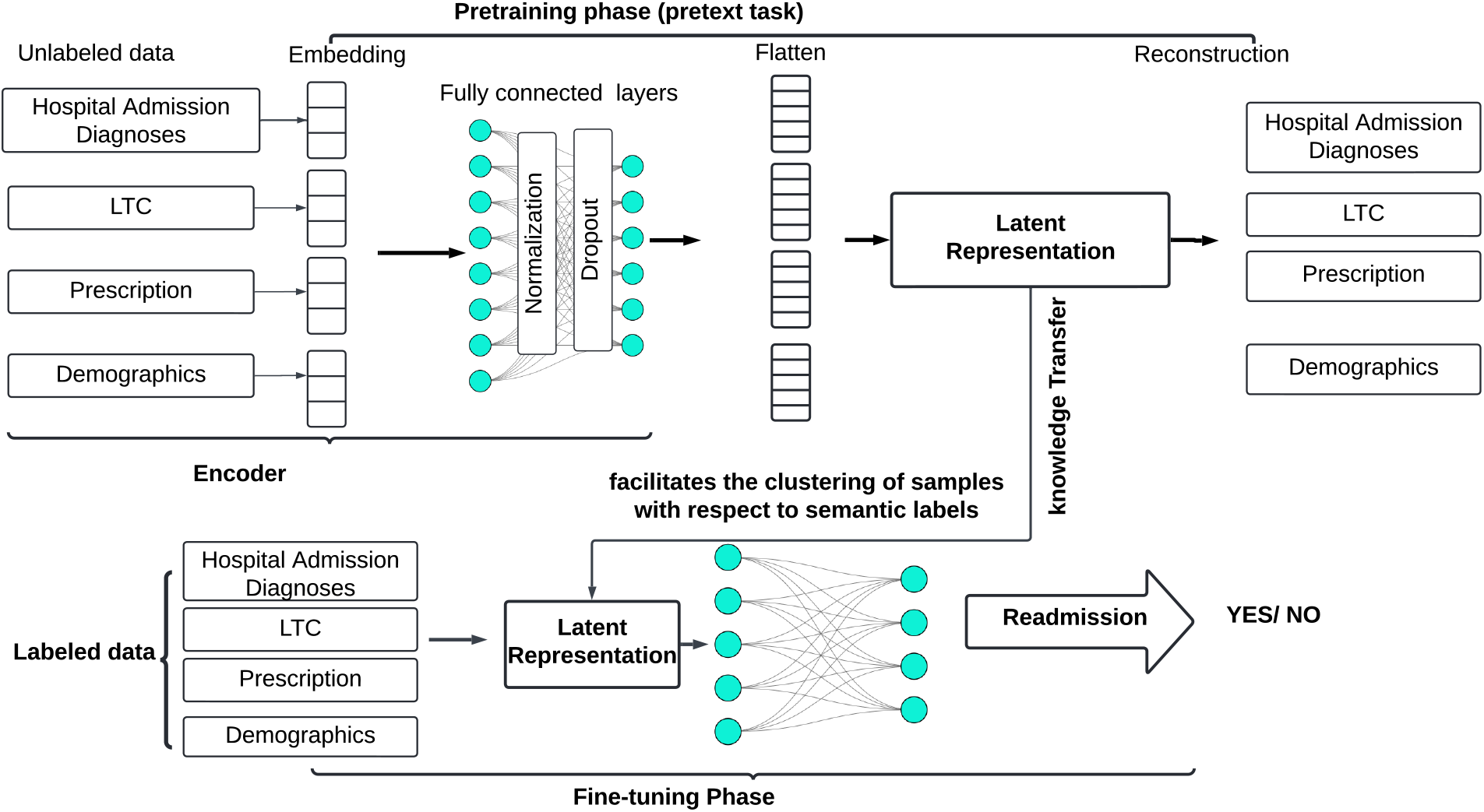
The Self-HR framework consists of two stages: self-supervised pre-training via an autoencoder (top), and supervised fine-tuning for hospital readmission (HR) classification (bottom). See text for a step-by-step explanation. Unlabelled data refers to lack of outcome variable (HR). LTC: long-term condition

Pre-training the model on self-supervised tasks also mitigates overfitting, a common issue when learning from imbalanced labelled datasets. In our model, Self-HR, an autoencoder is used for self-supervised pre-training. The autoencoder encodes each input into a compressed latent representation (embedding) and reconstructs the input from this representation by minimising reconstruction loss [22–24]. This pre-training serves as an effective form of regularisation by structuring the latent space, which ultimately improves performance on the downstream classification task. After pretraining, the model is fine-tuned using outcome labels (HR status) to perform binary classification. This two-stage pipeline (named Self-HR) is illustrated in Figure 3 and described below. For further details please see the supplementary material 3.

#### Pre-training Phase (Self-supervised)

1. Binary-encoded vectors for ICD-10, BNF, and LTC inputs are passed through feature-group–specific dense layers to obtain 50-dimensional embedded representations.
2. Each group passes through a dense layer, batch normalisation, and dropout.
3. Embedded representations are concatenated and compressed into a joint latent space (bottleneck).
4. The decoder reconstructs the original inputs from this latent vector.
5. Training minimises MSE reconstruction loss until convergence.

#### Fine-tuning Phase (Supervised)

1. Bottleneck representations are passed through a dense layer and dropout.
2. A final dense layer with sigmoid activation performs binary classification.
3. Binary cross-entropy loss is used for training.

### 2.7 Fully Supervised Baseline Models

To benchmark Self-HR, four fully supervised models were trained on the complete labelled dataset: a Random Forest (RF), XGBoost, a simple neural network (Simple NN), and a complex neural network (Complex NN).

**Random Forest and XGBoost** are ensemble methods well suited to tabular clinical data. RF constructs multiple decision trees using bootstrap aggregation and averages their predictions to reduce variance [25]. XGBoost extends gradient boosting with regularisation and second-order gradient optimisation, offering strong performance on structured EHR data [26]. Hyperparameter optimisation for both models is described in Supplementary Section 4.

**Simple NN** takes all input feature groups concatenated into a single vector, which is passed directly to a hidden layer followed by a sigmoid output unit for binary classification, without any feature-group-specific transformation layers (Figure 4, right).

**Fig. 4:**
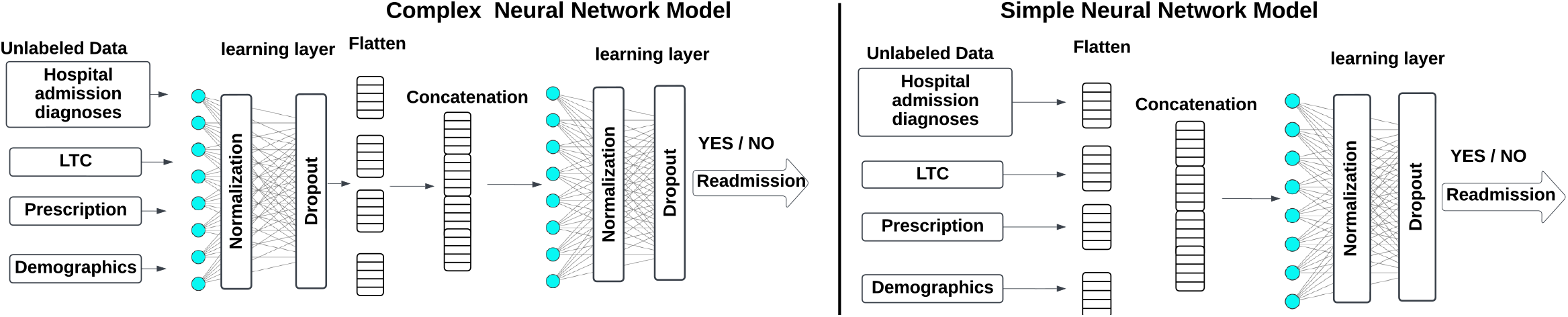
Neural network architectures: Complex model (left) and Simple model (right). LTC: long-term condition.

**Complex NN** is a multi-layer perceptron (MLP) that extends the simple architecture by incorporating multiple fully connected transformation layers applied independently to each input group (ICD-10 diagnoses, BNF prescriptions, LTCs, and demographics) prior to concatenation (Figure 4, left). The outputs of these group-specific transformation layers are then concatenated and passed through a final set of fully connected layers for binary classification. This design allows the model to learn feature-group-specific representations before joint classification, more closely mirroring the Self-HR encoder structure whilst remaining fully supervised. Hyperparameter search spaces for both neural network architectures are provided in Supplementary Tables 5 and 6.

All supervised models were trained using binary cross-entropy loss with the Adam optimiser and evaluated under identical data splits and testing conditions as Self-HR to ensure fair comparison.

### 2.8 Hyperparameter optimisation

Final settings for network-specific hyperparameters were determined through an initial grid search and validation-based optimisation for the Self-HR. We explored learning rates from 0.0001 to 0.01, batch sizes of 32, 64, and 128, dropout rates ranging between 15% and 50%, and epochs from 20 to 50. We selected hyperparameters via grid search over predefined ranges using validation performance and early stopping.

From these experiments, a learning rate of 0.0001 was selected as it consistently yielded stable convergence, while higher rates (e.g., 0.001 or 0.01) caused oscillations or divergence. A batch size of 64 struck the best balance between computational efficiency and generalisation: smaller sizes (e.g., 32) produced noisier gradient updates, and larger sizes (e.g., 128) slowed convergence. For dropout, 20% effectively reduced overfitting without compromising model expressiveness; lower values (e.g., 15%) provided insufficient regularisation, and higher values (e.g., 30% or 50%) degraded performance by excessively restricting the network’s capacity. Batch normalisation was applied after each layer to stabilise training and accelerate convergence [27]. Early stopping was employed to dynamically determine the optimal number of epochs, halting training once validation loss stopped improving to prevent overfitting.

The embedding dimension was chosen as 50 through grid search during pretraining, balancing the trade-off between minimising reconstruction loss and maintaining a compact latent representation.

For baseline RF models, we initially used scikit-learn’s default of 100 trees and validated this choice by testing 50, 100, and 200 trees. Although 200 trees marginally improved performance, the increased computational cost outweighed the benefits. Hyperparameter optimisation for the XGBoost model was performed using a grid search over commonly used parameter ranges for tabular clinical prediction tasks. The explored parameter ranges are summarised in Supplementary Table 4. NN architectures were initialised using heuristics [28], with the number of hidden units set to half the input layer size. We refined these values via incremental adjustments (doubling or halving) and tested architectures with 1 to 3 hidden layers. Final configurations were selected based on superior validation performance. Detailed hyperparameter selections are presented in Supplementary Tables 5 and 6.

### 2.9 Evaluation metrics

We evaluated Self-HR performance using AUROC, AUPRC, and F1 score. Both AUROC (Area Under the ROC Curve) and AUPRC (Area Under the Precision-Recall Curve) were reported due to the imbalanced nature of our data. Additionally, The F1 score balances precision and recall, offering a more informative metric for imbalanced datasets than precision or recall alone [29].

### 2.10 Ablation Study

To evaluate the contribution of different input feature groups to predictive performance, we selected the Random Forest (RF) as a baseline comparator for the proposed Self-HR model. We conducted a comprehensive ablation study in UK Biobank participants, assessing the performance of both Self-HR and RF while systematically excluding one feature group at a time (prescriptions, hospital admission diagnoses, demographic variables, and LTC characteristics) while retaining all remaining inputs.

### 2.11 Patient and public involvement and engagement

This research is part of a larger project, AI-MULTIPLY (ai-multiply.co.uk), which works closely with the Patient and Public Involvement and Engagement (PPIE) group. Two patients from our PPIE group with lived experience of MLTCs attended data engineering meetings while our methodology was being formulated, refined and later when the results were discussed. Their feedback was assimilated into our analysis plans and reporting of the results.

## 3 Results

### 3.1 Patient Cohort and Characteristics

Demographic and clinical characteristics of both the discovery (UKBB) and replication (CPRD) datasets are summarised by ethnic group in Table 1.

**Table 1:** Demographic and Clinical Characteristics by Ethnic Group in the UK Biobank and CPRD Datasets.

| **UK Biobank Dataset** |  |  |  |  |  |  |  |
| --- | --- | --- | --- | --- | --- | --- | --- |
| Ethnic Group | White | Asian or Asian British | Black or Black British | Chinese or Other Group | Mixed | Unknown | Total |
| Total | 94123 (94.12%) | 2136 (2.15%) | 1112 (1.12%) | 842 (0.85%) | 473 (0.48%) | 521 (0.53%) | 99207 (100%) |
| Sex |  |  |  |  |  |  |  |
| Female | 52380 | 1034 | 729 | 515 | 311 | 236 | 55205 |
| Male | 41743 | 1102 | 383 | 327 | 162 | 285 | 43802 |
| Age at 2nd LTC |  |  |  |  |  |  |  |
| Median (Q1, Q3) | 49 (40, 56) | 47 (39, 54) | 43.5 (37, 51) | 46 (40, 53) | 43 (35, 50) | 47 (40, 55) |  |
| Age at Incident Admission |  |  |  |  |  |  |  |
| Median (Q1, Q3) | 65 (57, 72) | 61 (53, 69) | 56 (49, 65) | 59 (51, 67) | 57 (49, 64) | 63 (55, 70) |  |
| IMD Quintile (1 = most deprived) |  |  |  |  |  |  |  |
| Median (Q1, Q3) | 3 (2, 4) | 2 (1, 3) | 1 (1, 2) | 2 (1, 3) | 2 (1, 4) | 2 (1, 4) |  |
| Prescriptions* |  |  |  |  |  |  |  |
| Median (Q1, Q3) | 6 (4, 10) | 10 (5, 15) | 8 (4, 12) | 7 (4, 12) | 7 (4, 11) | 8 (4, 13) |  |
| LTCs** |  |  |  |  |  |  |  |
| Median (Q1, Q3) | 6 (4, 9) | 7 (4, 10) | 6 (3, 9) | 5 (3, 9) | 5 (3, 9) | 6 (4, 9) |  |
| **CPRD Dataset** |  |  |  |  |  |  |  |
| Ethnic Group | White | Asian or Asian British | Black or Black British | Chinese or Other Group | Mixed | Unknown | Total |
| Total | 69974 (88.34%) | 3253 (4.11%) | 2231 (2.82%) | 1679 (2.12%) | 520 (0.66%) | 1567 (1.98%) | 79224 |
| Sex |  |  |  |  |  |  |  |
| Female | 36119 | 1686 | 1281 | 851 | 298 | 782 | 41017 |
| Male | 33855 | 1567 | 950 | 828 | 222 | 785 | 38207 |
| Age at 2nd LTC |  |  |  |  |  |  |  |
| Median (Q1, Q3) | 44 (34, 52) | 46 (39, 53) | 44 (38, 51) | 47 (40, 54) | 43 (37, 50) | 45 (37, 54) |  |
| Age at Incident Admission |  |  |  |  |  |  |  |
| Median (Q1, Q3) | 56 (47, 64) | 55 (48, 63) | 51 (45, 59) | 55 (48, 62) | 52 (45, 60) | 56 (48, 65) |  |
| IMD Quintile |  |  |  |  |  |  |  |
| Median (Q1, Q3) | 3 (2, 4) | 3 (2, 4) | 2 (1, 3) | 3 (2, 4) | 3 (1.75, 4) | 3 (2, 4) |  |
| Prescriptions* |  |  |  |  |  |  |  |
| Median (Q1, Q3) | 7 (4, 13) | 11 (6, 19) | 8 (4, 14) | 8 (4, 14) | 8 (4, 13) | 5 (3, 10) |  |
| LTCs** |  |  |  |  |  |  |  |
| Median (Q1, Q3) | 4 (3, 7) | 5 (3, 8) | 4 (2, 6) | 4 (3, 6) | 4 (2.75, 6) | 3 (2, 5) |  |

\*Counts of unique BNF paragraphs in 2 years prior to incident admission
\*\*Counts of distinct LTCs prior to incident admission
BNF: British National Formulary, CPRD: Clinical Practice Research Datalink, IMD: Index of Multiple Deprivation, LTC: Long Term Condition.

#### 1. UK Biobank discovery dataset

Of 502,486 volunteer participants, a cohort of 99,207 participants (ages 40–69 years) met our defined inclusion criteria (Figure 1). Of these participants, 14, 276 (14.4%) were assigned a positive readmission status (emergency readmission within 30 days of any previous discharge, based on admission method codes defined in the NHS Data Model and Dictionary). Most patients were of white ethnicity (94.1%) and female (55.6%) (Table 1). Asian or Asian British participants (2.2%) overall had the highest number of median LTCs (n=7) and prescriptions (n=10) versus all other ethnic groups (n=6 for LTCs and prescriptions in white population). Compared to all other ethnic groups, median age at second LTC diagnosis was lowest for those with mixed ethnicity (age 43, versus age 49 for white population). Median age at incident admission and death was lowest for participants of Black or Black British ethnicity (1.12%, median age at incident admission: 56, death: 66).

#### 2. Clinical Practice Research Datalink (CPRD) replication dataset

Of the 2, 677, 411 participants registered to CPRD between 2006 and 2010, 168,374 participants met our specified inclusion criteria (Figure 1). A cohort of 20, 120 (25.4%) were assigned a positive readmission status. Most patients were of white ethnicity (88.3%) (Table 1). Black or Black British participants had lowest median age (51 years) at incident admission across ethnic groups (56 years for white population). The mixed ethnicity group had the lowest median age at death (58 years, versus 68 years for white population).

As expected, the CPRD cohort demonstrated greater ethnic diversity than UKBB, with a higher proportion of participants from non-white ethnic groups [30] (Table 1). Sex distribution was more balanced in CPRD relative to UKBB. Median age at second LTC was highest among White participants (49 [40, 56]), while a higher relative number of LTCs was observed in Asian or Asian British individuals (median 7 [4, 10]) compared with other ethnic groups.

### 3.2 Model performance

All models demonstrated meaningful predictive capability in this multimorbid population, with AUROC values consistently exceeding 0.85. Table 2 reports performance across all five models on both the UKBB and CPRD cohorts under original class proportions, while Table 3 isolates the Simple NN and Self-HR models to examine how performance degrades under reduced label availability — a setting that directly probes the practical value of self-supervised pre-training.

**Table 2:** Performance of HR predictions using original subject proportions from the UKBB (left) and CPRD (right) datasets across five models: XGBoost, RF, Simple Neural Network model, Complex NN model and Self-HR. AUROC: area under receiver operating characteristic curve, AUPRC: area under precision recall curve, NN: neural network

| UKBB |  |  |  |  |  |  | CPRD |  |  |  |  |
| --- | --- | --- | --- | --- | --- | --- | --- | --- | --- | --- | --- |
| Model | Readmission | Precision | Recall | F1 | AUROC | AUPRC | Precision | Recall | F1 | AUROC | AUPRC |
| XGBoost | Yes | 0.89 | 0.16 | 0.27 | 0.85 | 0.59 | 0.86 | 0.34 | 0.49 | 0.81 | 0.67 |
|  | No | 0.88 | 1.00 | 0.93 |  |  | 0.81 | 0.98 | 0.89 |  |  |
| Complex NN | Yes | 0.74 | 0.42 | 0.54 | 0.88 | 0.65 | 0.79 | 0.56 | 0.65 | 0.85 | 0.76 |
|  | No | 0.90 | 0.97 | 0.94 |  |  | 0.86 | 0.94 | 0.90 |  |  |
| Simple NN | Yes | 0.86 | 0.28 | 0.42 | 0.88 | 0.68 | 0.81 | 0.51 | 0.65 | **0.86** | **0.76** |
|  | No | 0.89 | 0.99 | 0.93 |  |  | 0.86 | 0.96 | 0.91 |  |  |
| RF | Yes | 0.81 | 0.28 | 0.42 | 0.86 | 0.62 | 0.87 | 0.38 | 0.53 | 0.84 | 0.73 |
|  | No | 0.89 | 0.88 | 0.94 |  |  | 0.82 | 0.98 | 0.89 |  |  |
| Self-HR | Yes | 0.82 | 0.75 | 0.72 | **0.92** | **0.75** | 0.81 | 0.53 | 0.67 | **0.86** | **0.76** |
|  | No | 0.95 | 0.92 | 0.94 |  |  | 0.86 | 0.96 | 0.90 |  |  |

**Table 3:** Performance of HR predictions using original subject proportions from UKBB (left) and CPRD (right) datasets for Simple NN and Self-HR models. Refer to table 4 for abbreviations

| Models | UKBB |  |  |  |  |  | CPRD |  |  |  |  |
| --- | --- | --- | --- | --- | --- | --- | --- | --- | --- | --- | --- |
|  | Readmission | Precision | Recall | F1 | AUROC | AUPRC | Precision | Recall | F1 | AUROC | AUPRC |
| Simple NN<br>100% labelled data | Yes | 0.86 | 0.28 | 0.42 | 0.88 | 0.68 | 0.81 | 0.51 | 0.65 | 0.86 | 0.76 |
|  | No | 0.89 | 0.99 | 0.93 |  |  | 0.86 | 0.96 | 0.91 |  |  |
| Simple NN<br>50% labelled data | Yes | 0.43 | 0.20 | 0.28 | 0.85 | 0.43 | 0.76 | 0.41 | 0.53 | 0.79 | 0.65 |
|  | No | 0.87 | 0.96 | 0.91 |  |  | 0.83 | 0.96 | 0.89 |  |  |
| Self-HR 100%<br>labelled data | Yes | 0.82 | 0.75 | 0.72 | **0.92** | **0.75** | 0.81 | 0.53 | 0.67 | 0.86 | 0.76 |
|  | No | 0.95 | 0.92 | 0.94 |  |  | 0.86 | 0.96 | 0.90 |  |  |
| Self-HR 50%<br>labelled data | Yes | 0.50 | 0.83 | 0.62 | 0.91 | 0.72 | 0.76 | 0.53 | 0.33 | 0.84 | 0.74 |
|  | No | 0.96 | 0.86 | 0.91 |  |  | 0.85 | 0.95 | 0.90 |  |  |

In the UKBB cohort, Self-HR consistently outperformed all supervised models for predicting unplanned hospital readmission (HR) across multiple evaluation metrics, including F1 score for both the readmitted (0.72) and non-readmitted (0.94) classes, AUROC (0.92), and AUPRC (0.75), as shown in Table 2. The performance advantage of Self-HR is particularly evident when compared with traditional ensemble methods and fully supervised neural networks. XGBoost and RF achieved substantially lower F1 scores for the readmitted class (0.27 and 0.42, respectively), while recall for the clinically critical minority class was markedly higher for Self-HR (0.75) than for XGBoost (0.16) and the Simple NN (0.28).

Consistent performance trends were observed in the replication CPRD cohort, where Self-HR maintained strong predictive accuracy (AUROC 0.86, F1 score 0.67 for the readmitted class), despite a modest reduction relative to UKBB performance. Although AUROC was equivalent between Self-HR and the Simple NN on CPRD (both 0.86), Self-HR demonstrated meaningfully superior performance on the clinically more relevant minority class, achieving a higher F1 score for readmitted patients (0.67 vs. 0.65) and greater recall (0.53 vs. 0.51), indicating better sensitivity for identifying patients at genuine risk of readmission.

A key strength of the self-supervised learning paradigm is its ability to leverage unlabelled data during pre-training, reducing dependence on costly clinical annotations. Table 3 illustrates this directly by comparing Self-HR and the Simple NN under full (100%) and reduced (50%) label availability. Under full supervision, the Simple NN achieves AUROC values of 0.88 (UKBB) and 0.86 (CPRD); when label availability is halved, performance drops substantially to 0.85 and 0.79, respectively, with AUPRC falling from 0.68 to 0.43 on UKBB — a particularly pronounced degradation for the minority class. Self-HR, by contrast, proves considerably more resilient: with only 50% of labelled data, it retains an AUROC of 0.91 (UKBB) and 0.84 (CPRD) with AUPRC of 0.72 and 0.74, respectively, closely approaching its own fully supervised performance. This label efficiency reflects the representational advantage conferred by self-supervised pre-training, which allows the model to learn generalisable patient representations from the full dataset before fine-tuning on a smaller labelled subset.

Taken together, these findings demonstrate the robustness and generalisability of the Self-HR framework across distinct real-world clinical datasets with differing population characteristics, and underscore the practical value of self-supervised learning in low-annotation clinical settings where the early identification of patients at high risk of readmission is of direct clinical importance.

### 3.3 Self-HR robustness under partial outcome labelling

To further evaluate model robustness and label efficiency, we conducted experiments under partial outcome labelling, where readmission outcomes were available for only a subset of patients during downstream supervised training. A simple neural network was included as a comparator due to its similar performance to the Self-HR model observed in Table 2. Specifically, labels were provided for 50% of the UKBB training cohort while preserving the original class imbalance. Under this setting, the performance of the simple fully supervised simple NN deteriorated substantially, with the F1 score decreasing from 0.42 to 0.28 (Table 5). In contrast, Self-HR maintained considerably stronger performance, exhibiting a more moderate reduction in F1 score from 0.72 to 0.62. This pattern was even more pronounced in the CPRD replication dataset (Table 5).

### 3.4 Self-HR performance under downstream class-balancing strategies

To address the pronounced class imbalance in the discovery cohort, we evaluated the performance of Self-HR and comparison models under multiple class-balancing strategies. Such balancing strategies (random undersampling, CSL and SMOTE) were applied exclusively during the downstream supervised classification stage, following self-supervised representation learning. (Table 4). Across all balancing strategies, recall for the readmitted class improved, reflecting enhanced sensitivity to clinically important events. Among the evaluated approaches, Self-HR with undersampling achieved the strongest overall performance, yielding the highest AUROC (0.93 ± 0.01) and AUPRC (0.76 ± 0.01), alongside favourable precision (0.65 ± 0.01) and F1-score (0.74 ± 0.01). Self-HR variants incorporating CSL and SMOTE also demonstrated consistently strong performance, achieving AUROC values of 0.86 ± 0.01 and 0.88 ± 0.01, respectively, with balanced trade-offs between precision and recall. In contrast, complex NN and simple NN models, although competitive under balanced training conditions, consistently underperformed relative to Self-HR across most evaluation metrics. Collectively, these findings suggest that self-supervised pretraining produces feature representations that are resilient to downstream class-balancing strategies and well suited to real-world clinical deployment, where class imbalance is common. Notably, improvements were consistently observed in AUPRC, which is more informative than AUROC for evaluating readmission risk in highly imbalanced clinical datasets.

**Table 4:** Mean ± standard deviation of HR prediction performance using shuffled training datasets in UKBB subjects with equal proportions of readmitted and non-readmitted subjects for: RF, Simple NN, Complex NN, and Self-HR. Results of Self-HR based on Synthetic Minority Oversampling Technique (SMOTE) and cost-sensitive learning (CSL) are also shown. Refer to table 4 for other abbreviations.

| Models | Readmission | Precision | Recall | F1 score | AUROC | AUPRC |
| --- | --- | --- | --- | --- | --- | --- |
| Simple NN +<br>undersampling | Yes | 0.66±0.03 | 0.45±0.01 | 0.54±0.03 | 0.87±0.03 | 0.62± 0.01 |
| | No | 0.91 ± 0.00 | 0.96±0.02 | 0.93±0.00 | | |
| RF +<br>undersampling | Yes | 0.55±0.02 | 0.84±0.02 | 0.66±0.00 | 0.86±0.00 | 0.70±0.05 |
| | No | 0.97±0.01 | 0.88±0.01 | 0.93±0.00 | | |
| Complex NN +<br>undersampling | Yes | 0.55±0.02 | 0.65±0.01 | 0.59±0.00 | 0.88±0.01 | 0.65±0.04 |
| | No | 0.94 ± 0.03 | 0.91±0.01 | 0.92±0.00 | | |
| Self-HR<br>based CSL | Yes | 0.72 ± 0.01 | 0.72±0.04 | 0.72±0.03 | 0.86±0.06 | 0.60±0.01 |
| | No | 0.95±0.00 | 0.95±0.00 | 0.96±0.01 | | |
| Self-HR<br>based SMOTE | Yes | 0.72±0.07 | 0.72±0.05 | 0.72±0.06 | 0.86±0.03 | 0.59±0.04 |
| | No | 0.95±0.01 | 0.95±0.04 | 0.96 ± 0.02 | | |
| Self-HR +<br>undersampling | Yes | 0.65±0.01 | 0.83±0.01 | 0.64±0.01 | **0.93±0.01** | **0.76±0.01** |
| | No | 0.96±0.00 | 0.87±0.01 | 0.91±0.00 | | |

### 3.5 Ablation Study

The ablation study results, presented in Table 5, demonstrate the relative importance of each feature group in predicting HR risk within the UKBB cohort.

**Table 5:** Ablation Study Results on HR Prediction Comparing Self-HR (left) and RF (right) Methods in UKBB subjects. LTC: Long-term condition. Refer to table 4 for other abbreviations.

| Self-HR Method |  |  |  |  |  |  | Random Forest (RF) |  |  |  |  |
| --- | --- | --- | --- | --- | --- | --- | --- | --- | --- | --- | --- |
| Input(s) | Readmission | Precision | Recall | F1 | AUROC | AUPRC | Precision | Recall | F1 | AUROC | AUPRC |
| With all inputs | Yes | 0.82 | 0.75 | 0.72 | **0.92** | **0.75** | 0.81 | 0.28 | 0.42 | 0.86 | 0.62 |
|  | No | 0.95 | 0.92 | 0.94 |  |  | 0.89 | 0.88 | 0.94 |  |  |
| Without Prescriptions | Yes | 0.78 | 0.32 | 0.45 | 0.87 | 0.62 | 0.56 | 0.86 | 0.68 | 0.87 | 0.72 |
|  | No | 0.89 | 0.98 | 0.93 |  |  | 0.97 | 0.89 | 0.93 |  |  |
| Without Hospital Admission Diagnoses | Yes | 0.51 | 0.38 | 0.17 | 0.67 | 0.28 | 0.46 | 0.81 | 0.59 | 0.82 | 0.64 |
|  | No | 0.86 | 0.99 | 0.92 |  |  | 0.96 | 0.84 | 0.90 |  |  |
| Without Demographics | Yes | 0.70 | 0.43 | 0.53 | 0.88 | 0.64 | 0.55 | 0.86 | 0.67 | 0.87 | 0.71 |
|  | No | 0.91 | 0.96 | 0.93 |  |  | 0.97 | 0.88 | 0.92 |  |  |
| Without LTC | Yes | 0.70 | 0.41 | 0.52 | 0.87 | 0.62 | 0.58 | 0.84 | 0.69 | 0.87 | 0.72 |
|  | No | 0.90 | 0.97 | 0.93 |  |  | 0.97 | 0.90 | 0.93 |  |  |

For the Self-HR model, the exclusion of hospital admission diagnoses resulted in the most substantial performance degradation. The AUPRC dropped from 0.75 to 0.28, while the F1-score for readmission cases decreased sharply from 0.72 to 0.17. These results highlight the critical importance of incident admission diagnostic information for accurate hospital readmission prediction. Primary care prescriptions within the last 2 years emerged as the second most influential feature category; their removal led to a 37.5% reduction in the F1-score for readmission cases (from 0.72 to 0.45) and a 17.3% decline in AUPRC (from 0.75 to 0.62). While demographics and LTC features showed comparable importance, the exclusion of LTC features had a slightly greater impact on model performance (AUPRC: 0.62 vs. 0.64 without demographics).

The RF model also showed a reduction in performance on removing hospital admission diagnosis. Removing these features caused a 22.6% decrease in AUPRC (from 0.86 to 0.64). Unlike Self-HR, the RF model showed better performance when prescription or LTC data were removed (AUPRC 0.72 vs. 0.62 with all inputs), and demonstrated greater relative dependence on LTC features, with their exclusion yielding marginally stronger performance than removing prescription data (AUPRC: 0.72 vs. 0.71). The RF model had superior recall for readmission cases when prescription data was excluded (0.86 compared to Self-HR’s 0.32), suggesting distinct feature utilisation patterns between the two models.

### 3.6 Cross-Task generalisation of Self-HR Representations

To evaluate the cross-task utility of the Self-HR encoder, a series of regression tasks were carried out using model fine-tuning alone, without retraining the pretrained encoder. Predicting hospital admission duration is clinically important for bed capacity planning, discharge coordination, and early identification of patients at risk of prolonged stays, particularly in MLTC populations where care complexity often drives extended hospitalisations. Accurate duration estimates could support more proactive discharge planning and more efficient allocation of inpatient resources. Table 6 demonstrates that majority of admission durations are of 1-2 days duration (median 2 days) with a positive skew. For incident admission duration, Self-HR achieved the lowest error (MAE 3.48, RMSE 8.62), with the log-transformed variant further reducing MAE to 3.00 (Table 7). This indicates improved modelling of the large proportion of short stays, though the accompanying increase in RMSE (9.05) reflects reduced sensitivity to long-stay outliers. A similar pattern was observed for emergency readmission duration, where Self-HR achieved superior performance with MAE 1.21 and RMSE 6.09, while the log-transformed model improved MAE to 1.14 but slightly increased RMSE to 6.15. For a new category of events, first emergency admission (with or without a previous incident admission), Self-HR again provided the strongest results (MAE 2.54, RMSE 8.34), with the log-transformed variant improving MAE to 2.45 but increasing RMSE to 8.45. Across all comparisons, both XGBoost and RF underperformed relative to Self-HR, even when trained on encoder-derived features. This confirms that endto-end fine-tuning of the self-supervised encoder provides a consistent and substantial advantage over using fixed representations.

**Table 6:** Distribution characteristics and duration-range breakdown for the three hospital admission duration outcomes.

| Task | Count (days) | Mean | Median | Std | Min | Max |
| --- | --- | --- | --- | --- | --- | --- |
| Incident Admission Duration | 98058 | 4.36 | 2 | 10.43 | 1 | 981 |
| Emergency Readmission Duration | 14035 | 7.43 | 4 | 14.95 | 1 | 414 |
| First Emergency Admission Duration | 65426 | 4.98 | 2 | 12.00 | 1 | 1147 |

| Days |  |  |  |  |  |  |  |  |
| --- | --- | --- | --- | --- | --- | --- | --- | --- |
| Task | 1–2 d | 3–5 d | 6–10 d | 11–25 d | 26–50 d | 51–100 d | 101–500 d | >500 d |
| Incident (%) | 59.1 | 26.2 | 12.3 | 5.8 | 1.4 | 0.4 | 0.2 | 0.0 |
| Emergency (%) | 38.8 | 25.8 | 18.1 | 12.3 | 3.5 | 1.1 | 0.4 | 0.0 |
| First Emergency (%) | 51.4 | 24.4 | 13.6 | 6.9 | 1.6 | 0.5 | 0.2 | 0.0 |

**Table 7:** Regression performance for predicting emergency and incident admission duration across different modelling approaches. Lower values indicate better performance. SSL: Self-Supervised Learning, RMSE: Root Mean Squared Error, Mean Absolute Error

| Task | Model Variant | RMSE (days) | MAE (days) |
| --- | --- | --- | --- |
| Incident Admission Duration | Self-HR (SSL raw) | **8.61** | **3.47** |
|  | Self-HR (SSL log) | 9.05 | 3.00 |
|  | XGBoost (raw) | 11.11 | 3.71 |
|  | XGBoost (enc.) | 11.18 | 3.67 |
|  | RF (raw) | 11.35 | 3.64 |
|  | RF (enc.) | 11.64 | 4.14 |
| Emergency Readmission Duration | Self-HR (SSL raw) | **6.09** | **1.21** |
|  | Self-HR (SSL log) | 6.15 | 1.14 |
|  | XGBoost (raw) | 6.87 | 1.60 |
|  | XGBoost (enc.) | 6.74 | 1.50 |
|  | RF (raw) | 6.97 | 1.51 |
|  | RF (enc.) | 6.84 | 1.70 |
| First Emergency Admission Duration | Self-HR (SSL raw) | **8.34** | **2.54** |
|  | Self-HR (SSL log) | 8.45 | 2.45 |
|  | XGBoost (raw) | 8.75 | 3.43 |
|  | XGBoost (enc.) | 8.82 | 3.24 |
|  | RF (raw) | 9.21 | 3.16 |
|  | RF (enc.) | 9.36 | 3.63 |

Note: “enc.” denotes encoder-based features of the Self-HR; “raw” denotes original input features.

In every task, log transformation improved MAE but increased RMSE, indicating a systematic trade-off: enhanced accuracy for common short stays at the expense of reduced robustness to less frequent prolonged admissions. The behaviour of log-transformed models is consistent with the heavy-tailed nature of hospital stay distributions. Log scaling increases sensitivity to short-duration cases but compresses the influence of long stays, explaining the observed MAE improvements alongside higher RMSE values. This distinction is clinically relevant as depending on whether a deployment prioritises accurate prediction of common cases or robust handling of long admissions, different transformations may be preferable.

## 4 Discussion

In this study, we have developed and validated Self-HR, a two-staged SSL framework for predicting unplanned HR across multimorbid populations. We show that the principal strength of this SSL approach lies in its ability to construct and learn individualised latent representations, or embeddings, that summarise a patient’s longitudinal medical history up to the point of incident admission. In the context of multimorbidity, where complex and interdependent relationships exist between diagnoses, medications, and demographic factors, the ability to learn such rich latent representations is particularly valuable.

Compared with standard machine and deep learning approaches, Self-HR enables accurate prediction under conditions of partial outcome labelling (HR status), supporting its translation into clinical settings where fully annotated training data cannot be assumed. In real-world observational datasets, severe class imbalance is common, either reflecting the true outcome distribution or arising from systematic biases in data collection. Such imbalance can impair a models ability to learn fair and generalisable representations, often leading to favouring of the majority class. Self-HR mitigates this challenge by leveraging self-supervised pretraining followed by fine-tuning on smaller labelled datasets, enabling the identification of subtle but clinically meaningful risk signals even in highly imbalanced settings such as HR prediction. Nonetheless, we show that performance may be further improved through the incorporation of class-balancing strategies.

Our ablation study provided strong empirical support for the inclusion of diagnostic data of incident admission together with primary care prescription and LTC information. This is of particular relevance in the context of multimorbidity, where MLTCs are a risk factor for readmission [6]. These ablation study findings additionally align with established clinical understanding, whereby readmissions frequently arise from reoccurrence or progression of the primary presenting condition, and HR risk varies substantially according to the patients index diagnosis [31].

Building on this, a further key strength and motivation for adopting a SSL approach is its capacity for cross-task generalisability. The Self-HR encoder, pretrained to learn task-agnostic representations, can be fine-tuned for both readmission classification and related regression tasks, including prediction of durations of incident hospital admission and unplanned readmissions with 30 days. This reduces reliance on large, fully labelled datasets and avoids the need for repeated model redevelopment across clinical prediction problems.

Consistent with this, Self-HR more effectively captured the complexity of hospital admission durations than supervised models, with improved MAE and RMSE across all regression tasks, indicating enhanced prediction of both typical and prolonged admissions. Across tasks, RMSE values remained substantially higher than MAE, reflecting the skewed and heavy-tailed distribution of admission durations. Notably, Self-HR achieved the lowest RMSE in each task, suggesting improved handling of less common, but clinically important long-stay cases. Self-HR also demonstrated strong performance in predicting the duration of emergency admissions, where available clinical history may be limited. This suggests that in the case of MLTCs and polypharmacy, self-supervised representations learned from broader longitudinal contexts remain transferable and informative for such downstream tasks with constrained inputs. Collectively, these findings highlight the robustness of SSL for heterogeneous regression tasks in multimorbid populations and support suitability for real-world clinical deployment.

Self-HR outperforms many existing HR prediction models, which typically report AUROC values between 0.65-0.70[9, 32]. The modest performance of prior approaches is driven by heterogeneous readmission definitions, reliance on administrative data and the exclusion of clinically informative variables such as medication histories and longitudinal disease trajectories [5, 33]. While EHR data offer rich clinical information, they present challenges including missing data, coding variability, and high dimensionality, limiting the effectiveness of conventional modelling approaches [34].

The LACE index is a widely available tool predicting 30-day readmission or death using length of stay, admission acuity, comorbidity burden, and prior emergency visits. Although simple and interpretable, its predictive performance is limited (AUROC 0.60–0.68), restricting utility for individual-level clinical decision-making [35]. Machine learning models have demonstrated incremental improvements, with some achieving AUROC values above 0.70[36]. However, many focus on single-disease cohorts or older populations and often require extensive manual feature engineering[5].

Supervised deep learning approaches have sought to address these challenges by learning latent clinical patterns and temporal dependencies from EHR data. However, such approaches remain dependent on large, fully labelled datasets and have been developed for specific conditions or intensive care settings[37, 38]. In contrast, Self-HR can be adapted across multiple clinically relevant prediction tasks within a unified care pathway, supporting scalable, generalisable AI-driven risk stratification which could facilitate prioritisation of post-discharge interventions in patients with MLTCs.

## 5 Conclusions

People living with MLTCs experience complex care needs, high healthcare utilisation, and an elevated risk of unplanned 30-day hospital readmission. With the rising prevalence of multimorbidity, accurate readmission risk stratification is increasingly important to support targeted post-discharge interventions and the efficient allocation of healthcare resources. However, much of the existing readmission prediction literature remains focused on single-disease cohorts and fails to adequately capture the clinical heterogeneity, polypharmacy, and care complexity inherent to MLTC populations. To address this gap, we developed and externally validated Self-HR, a two-stage self-supervised learning framework that learns transferable patient representations from longitudinal clinical data encompassing diagnoses, prescriptions, long-term condition histories, and demographic factors. Self-HR achieved an AUROC of 0.92 and AUPRC of 0.75 in the UKBB discovery cohort, outperforming all supervised baselines across both overall and minority-class metrics, with external validation in 79,224 multimorbid CPRD participants confirming its generalisability. Crucially, Self-HR demonstrated superior robustness under partial outcome labelling and class imbalance — challenges that are endemic to real-world clinical datasets — maintaining an F1 score of 0.62 for readmitted patients even when trained on only 50% labelled data, compared with 0.28 for the best supervised comparator. Ablation analyses confirmed that incident admission diagnoses contributed most strongly to predictive performance, followed by primary care prescriptions and long-term condition history, findings that are consistent with established clinical understanding of readmission risk in multimorbid populations. Beyond binary classification, Self-HR demonstrated cross-task generalisability, achieving the lowest MAE and RMSE across three hospital admission duration regression tasks through fine-tuning alone, without any repeat pretraining. This highlights the potential of self-supervised representations to support a broader range of clinical prediction tasks within a unified framework, reducing the burden of model redevelopment across care pathway applications.

Collectively, these findings position Self-HR as a data-efficient, generalisable, and clinically grounded approach to readmission risk prediction in multimorbid populations. With further prospective validation, Self-HR has clear potential to support proactive discharge planning, targeted post-discharge interventions, and more equitable allocation of healthcare resources for people living with MLTCs.

## Acknowledgments

We are grateful to the UK Biobank participants. This research has been conducted using the UK Biobank resource under application number 69836. We would like to acknowledge the role of the AI-MULTIPLY Patient and Public Involvement and Engagement (PPIE) group, including Social Action for Health, throughout the project and the discussions leading to the work presented in this paper.

We would also like to thank the independent Advisory Board (IAB)—comprising Professor Frances Mair (Chair), Dr. Wasim Baqir, Dr. Claire Howard, and Professor Spiros Denaxas—for their rigorous strategic guidance and invaluable insights throughout the AI-MULTIPLY project. We extend our gratitude to Konstantin Shestopaloff and Zainab Awan for their critical review of the manuscript.

The views expressed are those of the authors and not necessarily those of the National Health Service, the NIHR or the Department of Health.

## Ethics

UK Biobank has received ethical approvals from the North West Multi-centre Research Ethics Committee (MREC), the Community Health Index Advisory Group (CHIAG), the Patient Information Advisory Group (PIAG) and the National Health Service National Research Ethics Service. Permission to use the UK Biobank resource for this research was approved by the UK Biobank Access Sub-Committee (approved research application no. 69836). The use of CPRD data for this study was approved by the Independent Scientific Advisory Committee for the Medicines and Healthcare products Regulatory Agency (reference 00068753). This study was conducted in accordance with the principles of the Declaration of Helsinki and all applicable UK ethical and regulatory requirements. UK Biobank participants provided written informed consent, and CPRD data were used in accordance with the CPRD governance framework following Independent Scientific Advisory Committee approval.

## Data availability

UK Biobank and The Clinical Practice Research Datalink (CPRD) do not allow the sharing of patient-level data. The data that support the findings of this study are available from UK Biobank (https://www.ukbiobank.ac.uk/about-our-data/) and CPRD (https://www.cprd.com/data), subject to registration and approval. The data were used under license for the current study.

## Materials availability

The code is available on GitHub at: https://github.com/rebeen/Readmission-Prediction-on-UK-BioBank-.git

## Author contribution

**Rebeen Ali Hamad**: Conceptualisation; Formal analysis; Data interpretation; Investigation; Methodology; Resources; Software; Validation; Visualisation; Roles/Writing Original Draft; and Writing review & editing. **Alisha Angdembe**: Conceptualisation; Data interpretation; Formal analysis. **Amaani B Hussain**: Data interpretation; Writing Original Draft. **Rafael Henkin**: Conceptualisation; Formal analysis. **John Casement**: Data curation; Resources; Software; Roles/Writing and Writing review & editing. **Wasim A Iqbal**: Conceptualisation; Data curation; Formal analysis; Investigation; Methodology; Writing review & editing. **Christian Atallah**: Conceptualisation; Formal analysis; Investigation; Methodology; Software. **Dexter Canoy**: Conceptualisation; Investigation; Methodology. **Rafel Henkin**: Conceptualisation; Formal analysis. **David Taylor**: Patient and Public group representative. **Susan Mountain**: Patient and Public group representative. **Michael Barnes**: Funding acquisition; Project leadership; Supervision; Data interpretation and review & editing. **Paolo Missier**: Conceptualisation; Funding acquisition; Project leadership; Supervision; Data interpretation; Roles/Writing original draft; and Writing review & editing. **Nick J Reynolds**: Conceptualisation; Funding acquisition; Project leadership; Supervision; Data interpretation; Roles/Writing original draft; and Writing review & editing.

### Clinical trial number

**not applicable**

## AI-MULTIPLY Consortium

Vanessa Apea^2,9^, Michael R. Barnes^2,10^, Victoria Bartle^6^, Alastair Burt^11^, Dexter Canoy^5^, Megan Clinch^12^, Ceri Durham^6^, Sarah Finer^13^, Olivia Grant^6^, Soraia Guerra-Sousa^5^, Barbara Hanratty^5^, John Isaacs^3,14^, Tom Lawton^15^, Hamish McAllister-Williams^11,16^, Paolo Missier^7^, Chris Plummer^14^, Nick J. Reynolds^3^, Sohan Seth^17,18^, Deborah Swinglehurst^12^, Adam Todd^19^, Neil Watson^14,20^, James Wason^21^

^9^ Barts Health NHS Trust, London, UK

^10^ Centre for Translational Bioinformatics, Queen Mary University of London, London, UK

^11^ Translational and Clinical Research Institute, Faculty of Medical Sciences, Newcastle University, Newcastle upon Tyne, UK

^12^ Centre for Public Health and Policy, Wolfson Institute of Population Health, Queen Mary University of London, London, UK

^13^ Blizard Institute, Barts and The London School of Medicine and Dentistry, Queen Mary University of London, London, UK

^14^ The Newcastle upon Tyne Hospitals NHS Foundation Trust, Newcastle upon Tyne, UK

^15^ Bradford Teaching Hospitals NHS Foundation Trust, Bradford, UK

^16^ Cumbria, Northumberland, Tyne and Wear NHS Foundation Trust, Newcastle upon Tyne, UK

^17^ School of Informatics, The University of Edinburgh, Edinburgh, UK

^18^ Advanced Care Research Centre (ACRC), The University of Edinburgh, Edinburgh, UK

^19^ School of Pharmacy, Faculty of Medical Sciences, Newcastle University, Newcastle upon Tyne, UK

^20^ Faculty of Medical Sciences, Newcastle University, Newcastle upon Tyne, UK

^21^ Population Health Sciences Institute, Faculty of Medical Sciences, Newcastle University, Newcastle upon Tyne, UK

